# Economic analyses of interventions for student mental health: protocol for a systematic review

**DOI:** 10.1101/2021.11.09.21255793

**Authors:** Eleanor Bell, Jia Pan, Chris Sampson

## Abstract

**Background:** Students in higher education exhibit higher rates of mental health problems than the general population and often lack access to effective treatments and services. The COVID-19 pandemic has exacerbated already high levels of need. Policymakers must have the relevant evidence to inform resource allocation and investment decisions about student mental health services.

**Aim:** We aim to identify and summarise economic analyses of interventions to prevent and treat mental health problems in university students in terms of the studies’ findings and the methods used.

**Method:** We will review all published economic analyses relating to interventions designed to prevent or treat student mental health problems. We will search the following databases: PubMed, MEDLINE, Embase, Web of Science, EconLit, PsycINFO and the National Health Service Economic Evaluation Databases (NHS EED). The review will be conducted following PRISMA guidelines. Key data items relating to the methods and results of the included studies will be extracted, and the resulting database will be made publicly available.

**Registration:** This review is registered on PROSPERO (ID CRD42021250228)

**Funding:** This research is funded by UK Research and Innovation (grant number ES/S00324X/1).

## Introduction

### Rationale

There is a growing body of research on the nature of mental health problems in university students in the UK (Gorczynski et al., 2017; Macaskill, 2013; Dodd et al., 2021; Stewart-Brown et al., 2000) and globally (Storrie, Ahern and Tuckett, 2010; Eisenberg, Golberstein and Gollust, 2007). Evidence shows that higher education students face significant unmet needs with respect to mental health, and service provision may be inadequate in many settings (Eisenberg, Golberstein and Gollust, 2007). Decision-makers in health care, public policy-making, and within universities require robust evidence to inform service provision and investment decisions.

Students exhibit higher rates of mental health problems compared to young adults who are not students (Cowan and Hao, 2021). Thus, there is a need to understand these high levels of prevalence and identify means of supporting students. The COVID-19 pandemic has brought this topic into sharp focus, with significant upheaval in higher education and the life of university students. These developments have given rise to novel research studies, demonstrating the impact of the pandemic on students and highlighting shortcomings in available services (Cameron, Fogarty-Perry and Piercy, 2021).

Previous reviews have explored the effectiveness of particular interventions for student mental health and wellbeing (Regehr, Glancy and Pitts, 2013; Farrer et al., 2013). However, no study to date (to our knowledge) has reviewed economic evidence for interventions and services to support student wellbeing. It is essential to identify economic evidence that can demonstrate the costs associated with an intervention, enabling decision-makers to compare alternative interventions and allocate resources appropriately.

A review of published evidence regarding the cost-effectiveness of interventions for student mental health and wellbeing may support the development of evidence-based programmes to improve student wellbeing. Furthermore, a review will inform methods for future research and identify gaps in the evidence, where further work could enhance services.

### Objective

This systematic literature review aims to identify and summarise the methods and findings of existing economic analyses of interventions to prevent and treat mental health problems in students.

## Methods and Analysis

This review will follow the Preferred Reporting Items for Systematic Reviews and Meta-Analyses (PRISMA) guidelines (Moher et al., 2009) and other relevant guidance for systematic reviews in this context (Khan et al., 2003; van Mastrigt et al., 2016).

### Eligibility criteria

The search will be guided by structured inclusion and exclusion criteria in the PICOS format (Khan et al., 2003). Comparative studies of interventions targetting student mental health, which report on economic evidence, will be eligible for inclusion, as described in detail in Table 1. We will exclude studies published in languages other than English; published earlier than January 2000; or for which the full study report is not available (i.e. excluding conference abstracts).

**Table 1:**
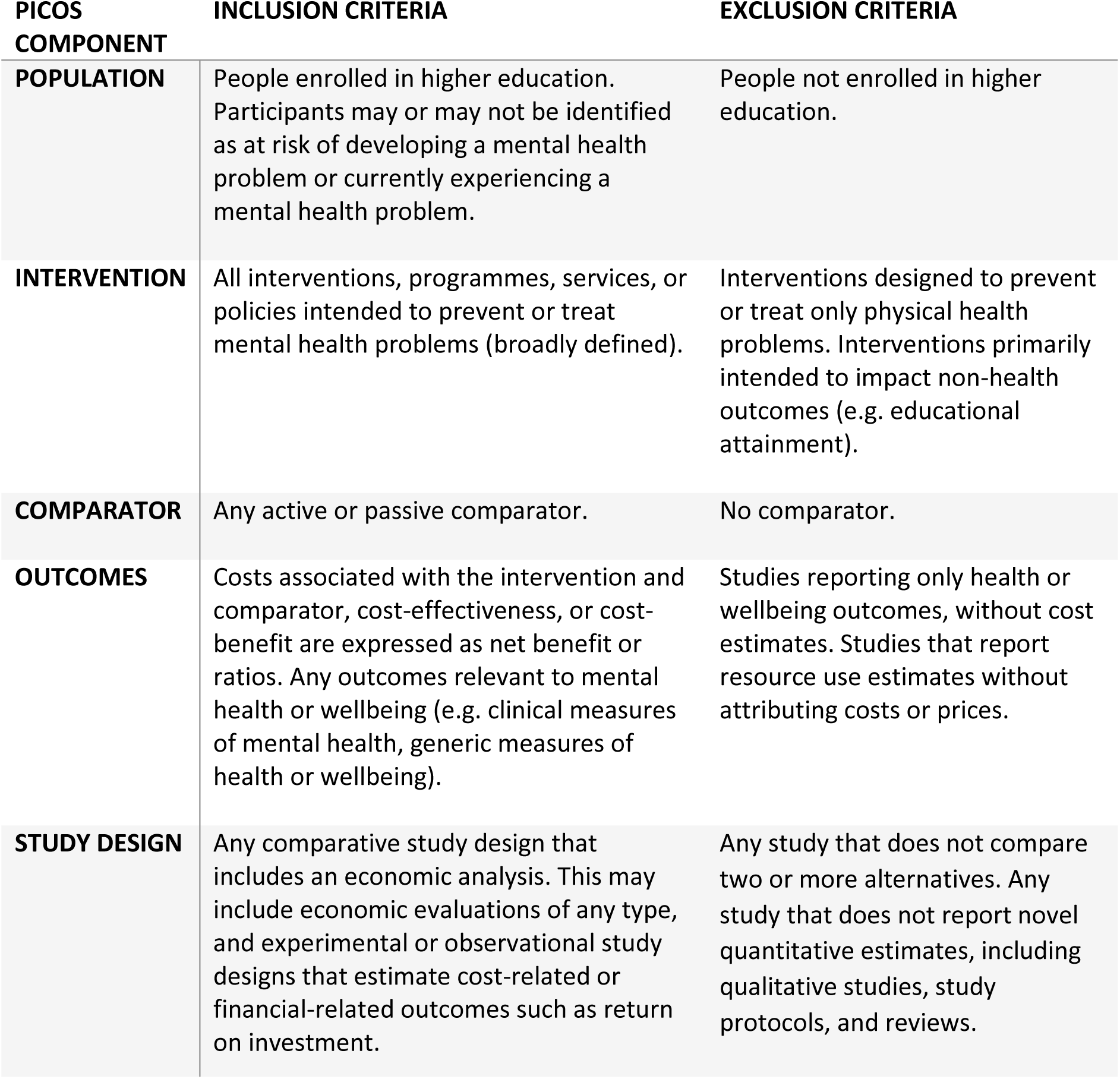
Eligibility criteria according to PICOS

| **PICOS COMPONENT** | **INCLUSION CRITERIA** | **EXCLUSION CRITERIA** |
| --- | --- | --- |
| **POPULATION** | People enrolled in higher education. Participants may or may not be identified as at risk of developing a mental health problem or currently experiencing a mental health problem. | People not enrolled in higher education. |
| **INTERVENTION** | All interventions, programmes, services, or policies intended to prevent or treat mental health problems (broadly defined). | Interventions designed to prevent or treat only physical health problems. Interventions primarily intended to impact non-health outcomes (e.g. educational attainment). |
| **COMPARATOR** | Any active or passive comparator. | No comparator. |
| **OUTCOMES** | Costs associated with the intervention and comparator, cost-effectiveness, or cost-benefit are expressed as net benefit or ratios. Any outcomes relevant to mental health or wellbeing (e.g. clinical measures of mental health, generic measures of health or wellbeing). | Studies reporting only health or wellbeing outcomes, without cost estimates. Studies that report resource use estimates without attributing costs or prices. |
| **STUDY DESIGN** | Any comparative study design that includes an economic analysis. This may include economic evaluations of any type, and experimental or observational study designs that estimate cost-related or financial-related outcomes such as return on investment. | Any study that does not compare two or more alternatives. Any study that does not report novel quantitative estimates, including qualitative studies, study protocols, and reviews. |

### Information sources and search strategy

We will search for literature in MEDLINE, Embase, Web of Science, EconLit, PsycINFO, and the National Health Service Economic Evaluation Databases (NHS EED).

We will use a combination of defined search words and search terms and Medical Subject Headings (MeSH) to identify studies of (1) interventions to prevent or treat mental health problems, (2) amongst students, which (3) report economic analyses. These search terms are specified with reference to previous review studies in the context of students or mental health (Donker et al., 2015; Farrer et al., 2013; Dodd et al., 2021; Fischer, Chwala and Simon, 2020; Mathes et al., 2014). An example search strategy is shown in Table 2.

**Table 2:**
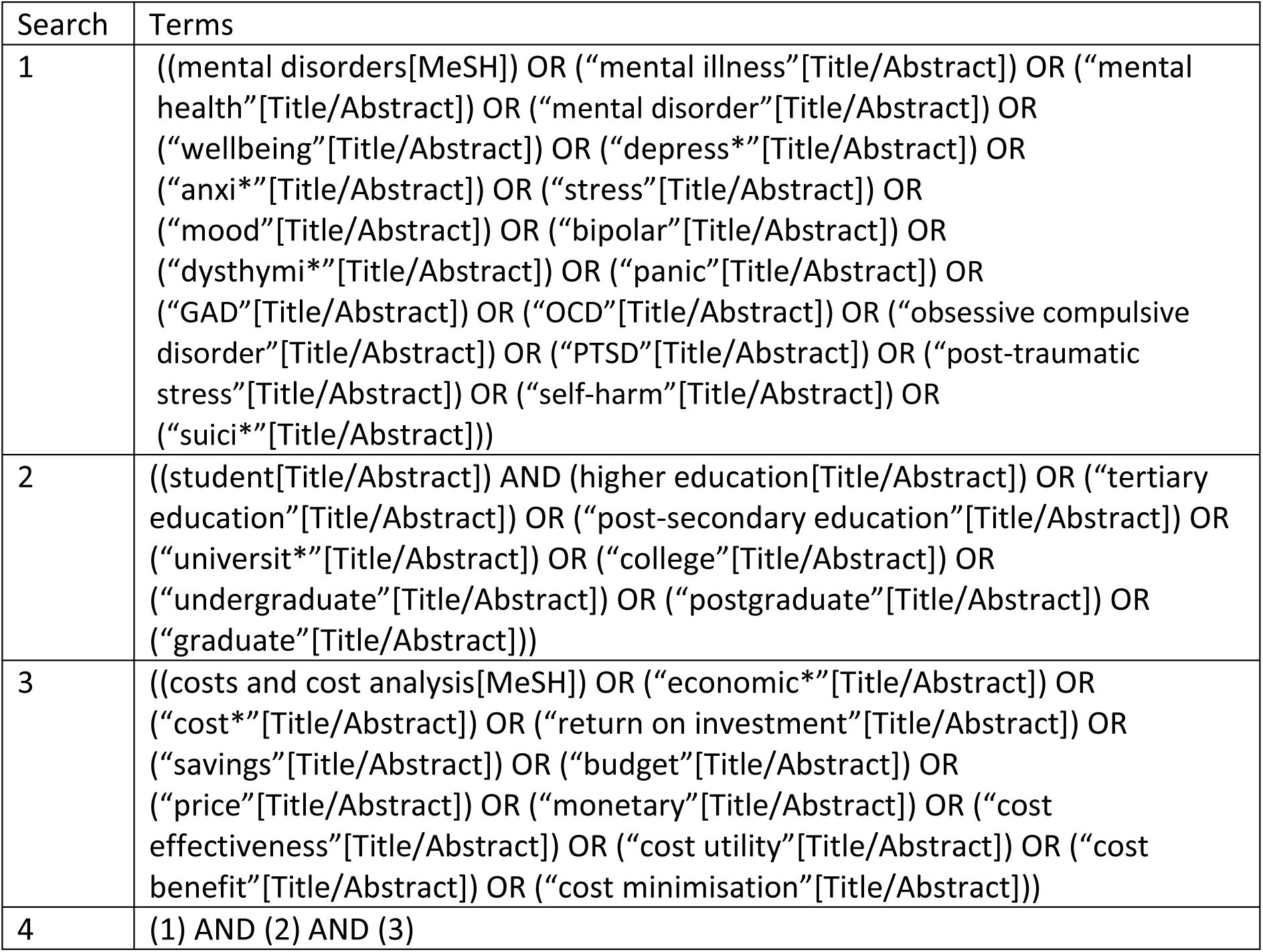
Example search strategy

We will scan references in any identified systematic reviews from our primary search to identify studies not previously identified.

### Study records

#### Data management

Titles and abstracts for identified citations will be recorded in a spreadsheet.

#### Selection process

The first reviewer will screen studies for retrieval based on title and abstract, according to the PICOS criteria. The second reviewer will check a random 20% of exclusions. If there are any disagreements, the second reviewer will check all excluded articles, and any disagreements will be resolved by discussion.

Full texts will be retrieved for studies not rejected at abstract screening. The first reviewer will assess these for satisfaction of the inclusion and exclusion criteria. Reasons for exclusion will be recorded and the second reviewer will check the validity of all exclusions. Any disagreements will be resolved by discussion. The number of records identified, retrieved, screened, assessed, included, and excluded in the review, and reasons for exclusions, will be summarised in a PRISMA flow diagram.

The reporting quality of included studies will be assessed using the CHEERS checklist (Husereau et al., 2013).

#### Data collection process

Data will be extracted by the first reviewer using an electronic data extraction form and automatically recorded in a spreadsheet. A second reviewer will subsequently cross-check extracted data for a random 20% of included studies. The following data items will be extracted:

∘ Article details
  - First author surname
  - Publication year
  - Study title
  - Publication name
  - DOI or URL
∘ Study characteristics
  - Study design (economic evaluation / other)
  - Comparators
  - Outcomes
  - Time horizon
∘ Sample characteristics
  - Sample size
  - Age range
  - Country
∘ Methods
  - Measures/questionnaires used
  - Data sources
∘ Results
  - Clinical/health outcomes
  - Costs
  - Resource use
  - Cost-effectiveness (if reported)

#### Data analysis and presentation

We will summarise the findings of all studies in both tabular and narrative overview formats. We will not pool estimates given that we anticipate variation in terms of interventions, comparators, and outcomes.

We anticipate a broad range of outcomes will be reported by included studies. These are likely to include the number of patients who responded to treatment; the number of participants in remission; changes in participants’ quality of life, reasons for dropping out of the study, and death. The primary resource use and cost outcomes will be changes in primary and secondary care resource use, and the monetary value. Cost-effectiveness outcomes are expected to be reported in ICERs. We will report all primary and secondary outcomes identified, grouped and categorised as appropriate.

There will be no attempt to conduct a meta-analysis, due to the anticipated heterogeneity of study designs and outcomes. We will not formally assess the quality of evidence as we expect there to be limited evidence of relatively low quality and our objective is to inform future research rather than service provision.

### Dissemination

Results will be submitted for publication in a peer-review journal and disseminated via the SmaRteN Student Mental Health Research Network. For transparency and reproducibility, the full database of literature that results from the data extraction will be published as an appendix to the systematic review.

## Discussion

To our knowledge, this will be the first systematic review conducted of economic analyses of student mental health interventions or services.

The research question we have adopted is broad, encompassing not only cost-effectiveness analysis studies but all economic analyses, encompassing all studies relating to the monetary impact of student mental health interventions. This systematic review will collate all currently available economic evidence in the literature and provide a comprehensive overview of the economic implications of student mental health interventions.

The evidence identified in this review may support policymakers and decision-makers in resource allocation decisions relating to the growing challenge of mental health in universities. The review will also provide an overview of methods used to date, which will guide future research.

## Data Availability

A database of the literature compiled as part of this systematic literature review will be made available for transparency.

